# Trends in Substance Use-related Emergency Department Visits by Youth, 2018-2023

**DOI:** 10.1101/2024.10.29.24316367

**Authors:** Madeline H. Renny, Yago Stecher, Carmen Vargas-Torres, Alexis M. Zebrowski, Roland C. Merchant

## Abstract

**Background:** Emergency departments (EDs) are a promising location for initiating substance use interventions for youth. Our objective was to determine trends in substance use-related ED visits for youth from 2018-2023, and investigate the patient characteristics, types of substance involved, and ED visit disposition and revisits.

**Methods:** We conducted a retrospective review of electronic health records (EHRs) from six EDs in an urban healthcare system to identify 12-21-year-old patients with a substance use-related ED visit from 2018 through 2023. Visits were identified by International Classification of Diseases, 10^th^ Revision Clinical Modification codes for substance use involving alcohol, cannabis, sedative/hypnotics, opioids, cocaine/stimulants, and psychoactive substances. The proportion of substance use-related visits each year was calculated by age group (12-14y, 15-17y, and 18-21y), sex, race/ethnicity, and substance type. We used 2-sample tests of binomial proportion to compare proportions. Logistic regression was used to assess characteristics associated with substance use-related visits, hospital admissions, and ED revisits.

**Results:** Of 151,764 ED visits for 12-21-year-olds, 4,556 (3.0%) were for substance use. From 2018-2023, substance use-related ED visits increased from 2.8% to 3.4% of all ED visits (p < 0.001) and were most often by 18-21-year-olds (79.4%), yet there were significant increases in visits by younger age groups (12-14y and 15-17y). Visits for females increased from 43.4% in 2018 to 52.4% in 2023 (p< 0.001). Although visits for alcohol were most frequent (53.1%), cannabis visits increased from 17.9% to 35.3%, with increases across all age groups (p < 0.001). Nineteen percent of visits involved patients that had an ED revisit for a substance use-related diagnosis within one year.

**Conclusion:** Substance use-related ED visits increased from 2018 to 2023, with an increase in visits for cannabis over time. These findings can inform targeted ED-based interventions for substance use in youth.

## 1. Introduction

Adolescent substance use is common and can lead to significant morbidity and mortality.^1,2^ Early initiation of substance use increases the risk for substance use disorder.^3,4^ Since the COVID-19 pandemic, there have been sustained lower rates of drug use for adolescents in the United States (US).^1^ Despite these trends, in 2023, there were still 29% of US 12^th^ graders who reported using cannabis, 46% using alcohol, and 7% using other illicit drugs within the prior year.^1^ Most concerning, drug overdose and poisoning are now the second leading cause of death for 15-24-year-olds in the US.^2,5^ These findings demonstrate the need to better understand youth substance use and opportunities for intervention to prevent addiction and death.

Among US adolescents, about 5% use the emergency department (ED) as their only source of health care.^6^ The ED visit therefore can be a critical access point for addressing the healthcare needs of youth with substance use. However, there is limited research describing ED visits for youth with substance use. Studies often report on combined substance use and mental health visits, which limits the ability to focus on the substance use subset of this population, or include all hospital visits, rather than focusing only on ED visits.^7-10^ A study analyzing National Hospital Ambulatory Medical Care Survey (NHAMCS) data on substance use and mental health visits by patients 11-24 years old from 1997 to 2010 found that substance use accounted for 2.1% of all ED visits, and that male gender, urban location, night and weekend shift times, and mental health disorders were associated with substance use visits.^7^ A more recent study investigated substance-related visits (both ED and hospitalizations) among youth at US children’s hospitals from 2016 to 2021, and observed an increase in visits for all ages, demographics, regions, and payors, but most notably for Hispanic youth and cannabis-related visits.^8^ Trends in substance use by adolescents have changed significantly over the past decade,^1^ and it is unknown if these changes have continued to impact substance use-related ED visits for adolescents across all age groups and different ED settings. Data on the most recent trends in the frequency of substance-use related visits, types of substances involved with these visits, and the outcomes of these visits, as well as how these trends vary by patient characteristics can inform the development of targeted ED-based interventions for youth. Therefore, the objective of this study was to determine the trends in substance-use related ED visits by youth in a multicenter healthcare system from 2018 to 2023, and compare these ED visits by patient characteristics, types of substance involved, ED visit disposition and revisits.

## 2. Methods

We conducted a retrospective review of electronic health records (EHRs) from six EDs in an urban healthcare system to identify 12-21-year-old patients with a substance-use related ED visit between January 2018 and December 2023. The six EDs care for pediatric patients and have a total annual pediatric patient volume of approximately 40,000 patients. One of these EDs admits patients to an adjoining children’s hospital. The study was approved by the Icahn School of Medicine at Mount Sinai Institutional Review Board.

### 2.1. Variables

The primary outcome of the study was the proportion of substance use-related ED visits (out of the total ED visits) for each year of the study. Substance use-related ED visits were defined as any visit associated with substance use (use, dependence, or overdose) and were identified using International Classification of Diseases, 10^th^ Revision Clinical Modification (ICD-10-CM) diagnosis codes.^11^ ICD-10-CM codes for substance use involving alcohol, cannabis, sedative/hypnotics, opioids, cocaine, stimulants, hallucinogens, inhalants, and other psychoactive substances were included (Supplementary Material, Table S1). Visits were classified by age group (12-14 years, 15-17 years, and 18-21 years) and substance type. Substance type was categorized as alcohol, cannabis, sedative/hypnotics, opioids, cocaine/stimulants, and psychoactive substances. The “psychoactive substances” category was based on ICD-10-CM codes for other psychoactive substances, as well as for hallucinogens and inhalants, due to the low number of ED visits for these substances.

### 2.2. Demographic Characteristics

Demographic variables related to the patient included age, sex (male or female), and race/Hispanic ethnicity (Non-Hispanic White, Non-Hispanic Black, Hispanic or Latino, Asian, and Other Non-Hispanic/Unspecified (including American Indian or Alaska Native/Native Hawaiian or Pacific Islander).

### 2.3. ED visit disposition and revisits

ED visit disposition was categorized as discharged home, admission (admission to hospital or admission to observation unit), transfer to psychiatry, or self-directed discharge (left without being seen, left before treatment complete, eloped, and/or left against medical advice). ED revisits were measured as repeat ED visits for substance use-related diagnoses within one week (day 2 to day 7) and within one year (day 2 to day 365) of the initial ED visit for a substance use-related diagnosis.

### 2.4. Analysis

The proportion of substance use-related ED visits was calculated as the number of substance use-related ED visits divided by the total number of ED visits. We calculated the proportion of substance use-related visits for each year of the study and then examined trends in substance use-related visits over time by age group, sex, and race/ethnicity. We used two-sample tests of binomial proportions and Chi-square two-tailed tests to compare substance use by demographic characteristics. We calculated the frequency of ED visit disposition and revisits for substance use-related visits for each year of the study period. We used logistic regression to assess changes in substance use-related visits over time and the demographic characteristics associated with substance use-related visits, admissions to the hospital, and ED revisits over time. Due to small sample size (n=2), we did not include visits for patients with a documented sex that was not male or female in the analysis. In our regression analysis, we used Hispanic as the reference category for race/Hispanic ethnicity, since Hispanic patients had the lowest proportion of substance use-related ED visits, and this would allow us to determine which patients were more likely to have a substance use-related visit. Analyses were performed using Stata/MP 17.0.

## 3. Results

### 3.1. Substance-use related ED visits and patient demographic characteristics

From 2018 through 2023, there were 151,764 visits by 12-21-year-old patients to the six EDs. Of these ED visits, 4,556 were substance use-related (3.0%). Substance use-related ED visits increased from 2.8% of all ED visits in 2018 to 3.4% in 2023 (Δ 0.62%, 95% CI 0.32% – 0.92%). Table 1 shows the demographic characteristics of patients presenting for substance use-related ED visits during each year of the study period. The majority of substance use-related visits (79.4%) were by youth 18-21 years old. The frequency of visits by males and females for a substance use-related visit were similar. The more often patients’ documented race/ethnicity was Other/Unspecified (30.4%) or Non-Hispanic White (25.2%). In the regression analyses, compared to 2018, there was a decreased odds of substance use-related visits in 2021 and an increased odds of substance use-related visits in 2022 and 2023 (Table 2). Male sex and older age were associated with increased odds of a substance use-related ED visit. As compared to Hispanic patients, patients with a documented race/ethnicity of Non-Hispanic White, Asian, and Other were more likely to have a substance use-related ED visit.

**Table 1.** Substance use-related emergency department visits for youth, 2018-2023.

|  | **2018** | **2019** | **2020** | **2021** | **2022** | **2023** | **Total** |
| --- | --- | --- | --- | --- | --- | --- | --- |
| **Total ED visits, n** | 23,071 | 25,990 | 17,829 | 28,108 | 28,818 | 27,948 | 151,764 |
| **Substance use-related ED visits, n (%)** | 643<br>(2.8) | 761<br>(2.9) | 502<br>(2.8) | 749<br>(2.7) | 950<br>(3.3) | 951<br>(3.4) | 4556<br>(3.0) |
| **Demographic characteristics of patients presenting for substance-use ED visits** |  |  |  |  |  |  |  |
| **Age group, n (%)** | **2018** | **2019** | **2020** | **2021** | **2022** | **2023** | **Total** |
| 12-14y | 8 (1.2) | 15 (2) | 21 (4.2) | 27 (3.6) | 38 (4) | 66 (6.9) | 175 (3.8) |
| 15-17y | 78<br>(12.1) | 118<br>(15.5) | 103<br>(20.5) | 113<br>(15.1) | 172<br>(18.1) | 179<br>(18.8) | 763 (16.7) |
| 18-21y | 557<br>(86.6) | 628<br>(82.5) | 378<br>(75.3) | 609<br>(81.3) | 740<br>(77.9) | 706<br>(74.2) | 3618 (79.4) |
| **Sex, n (%) \*** | **2018** | **2019** | **2020** | **2021** | **2022** | **2023** | **Total** |
| Female | 279<br>(43.4) | 350 (46) | 225<br>(44.8) | 400<br>(53.4) | 496<br>(52.2) | 498<br>(52.4) | 2248 (49.3) |
| Male | 364<br>(56.6) | 411 (54) | 277<br>(55.2) | 349<br>(46.6) | 454<br>(47.8) | 451<br>(47.4) | 2306 (50.6) |
| **Race/Hispanic ethnicity, n (%)** | **2018** | **2019** | **2020** | **2021** | **2022** | **2023** | **Total** |
| Non-Hispanic White | 188<br>(29.2) | 234<br>(30.8) | 121<br>(24.1) | 176<br>(23.5) | 216<br>(22.7) | 197<br>(20.7) | 1132 (24.8) |
| Non-Hispanic Black | 95<br>(14.8) | 133<br>(17.5) | 102<br>(20.3) | 138<br>(18.4) | 161<br>(17) | 175<br>(18.4) | 804 (17.6) |
| Hispanic or Latino | 151<br>(23.5) | 149<br>(19.6) | 92 (18.3) | 119<br>(15.9) | 173<br>(18.2) | 232<br>(24.4) | 916 (20.1) |
| Non-Hispanic Asian | 42 (6.5) | 58 (7.6) | 37 (7.4) | 49 (6.5) | 64<br>(6.7) | 54 (5.7) | 304 (6.7) |
| Other/unspecified | 167 (26) | 187<br>(24.6) | 150<br>(29.9) | 267<br>(35.7) | 336<br>(35.4) | 293<br>(30.8) | 1400 (30.7) |

\*2 indeterminate sex, unknown

**Table 2.**
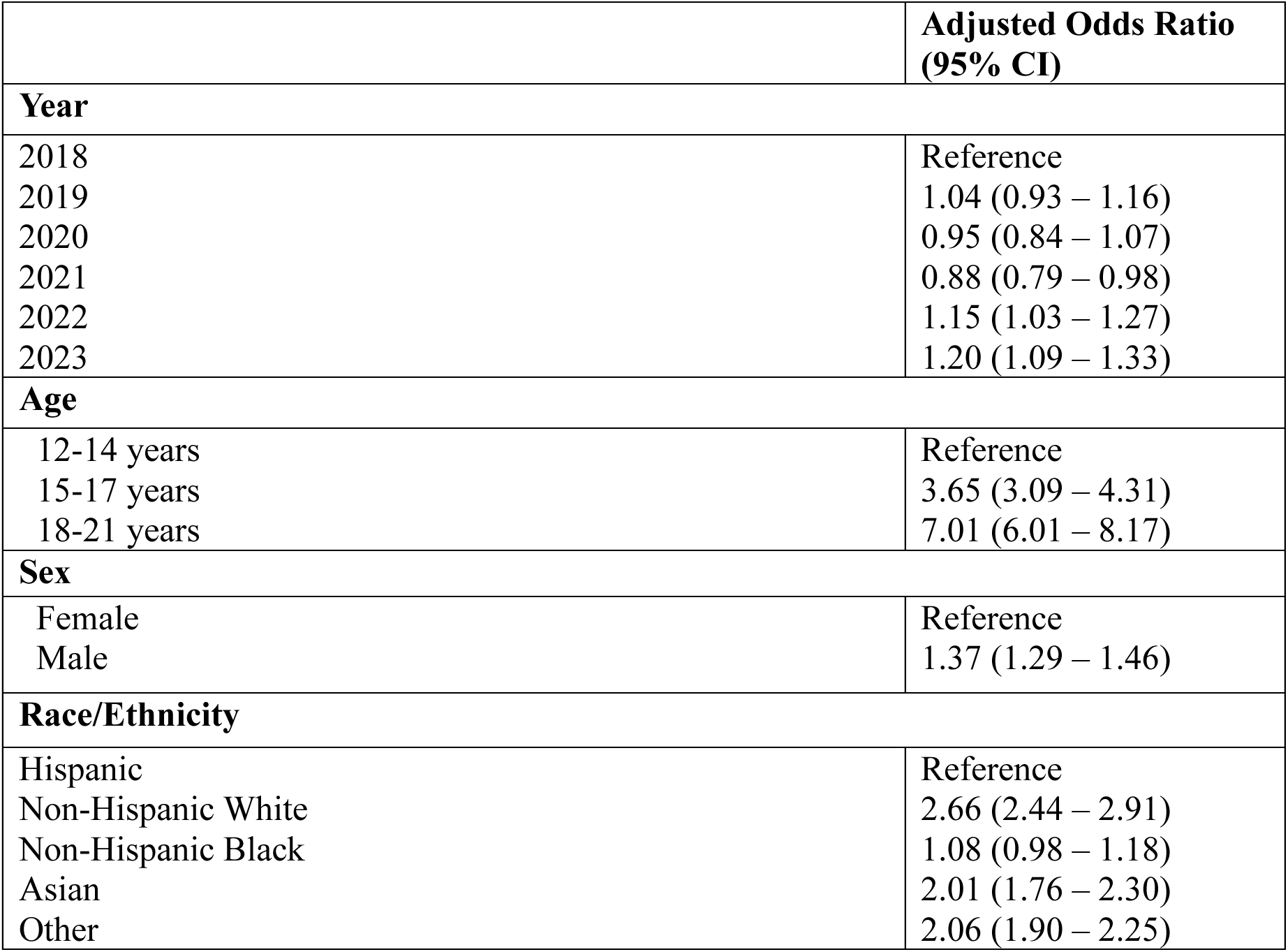
Factors associated with a substance use-related emergency department visit.

### 3.2. Age group trends in substance-use related visits

Trends in substance use-related visits varied over time and by age group (Figure 1). Between 2018 and 2023, the proportion of substance use-related ED visits for patients 12-14 years old increased from 0.17% to 1.17% (Δ 0.99%, 95% CI 0.69% – 1.30%). Visits for 15–17-year-olds increased from 1.40% to 2.64% (Δ 1.23%, 95% CI 0.74% – 1.72%). For both of these age groups, substance use-related visits initially increased, then decreased in 2021, with a subsequent increase in 2022 and 2023. The proportion of substance use-related visits for patients 18-21 years old were similar in 2018 and 2023, as the result of a decrease in 2020 and 2021, and a subsequent increase in 2022.

**Figure 1.**
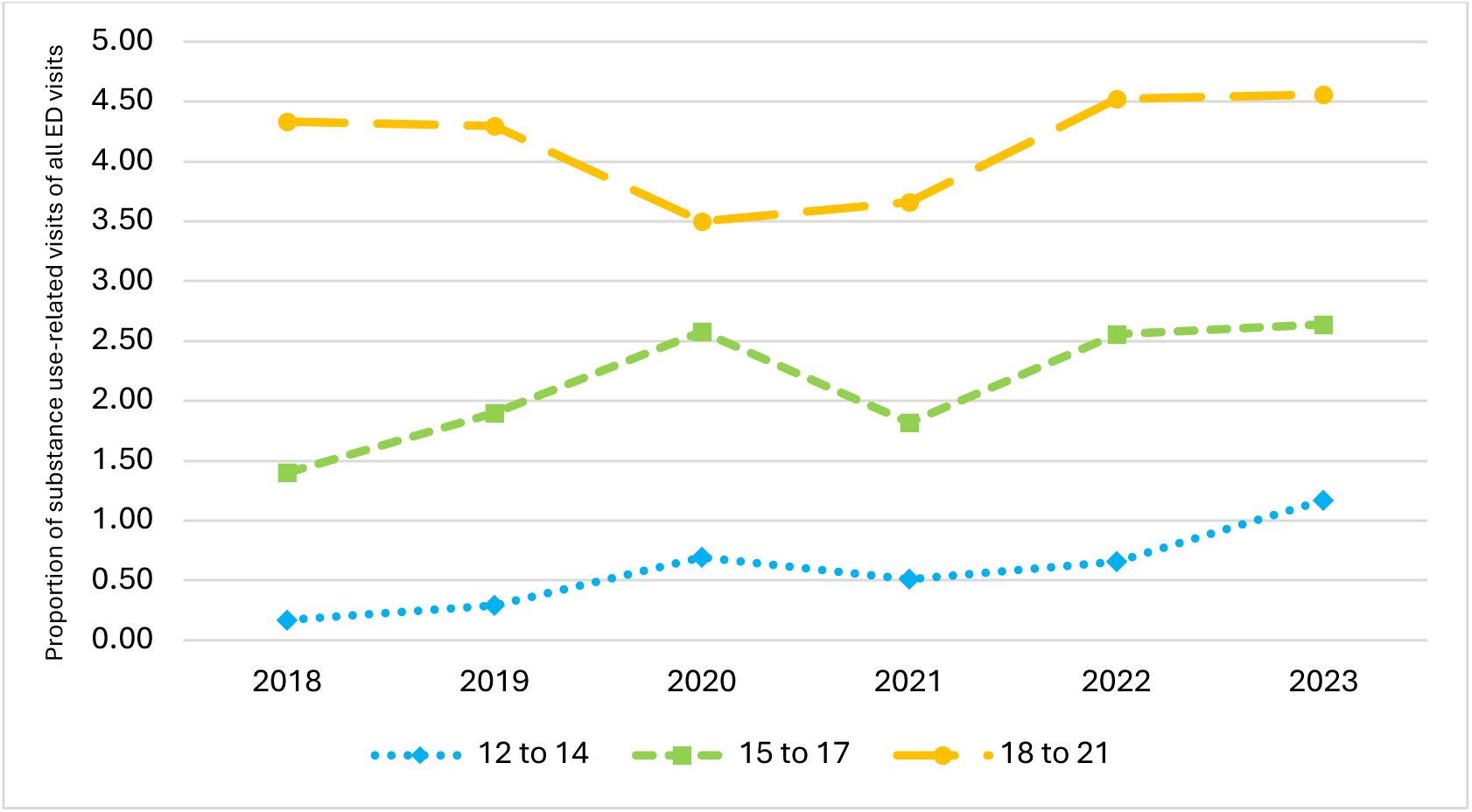
Substance use-related ED visits for youth by age group, 2018-2023.

### 3.3. Sex and race/Hispanic ethnicity trends in substance-use related visits

Trends in substance use-related visits varied by sex and race/Hispanic ethnicity (Figure 2a and 2b). In 2018, there were a greater proportion of substance use-related visits for males (56.6%), but the proportion of substance use-related visits for females increased during the study period, and by 2023, 52.3% of visits were for females. As shown in Figure 2a, the proportion of ED visits for females with substance use increased from 2.20% in 2018 to 3.20% in 2023 (Δ 1.00%, 95% CI 0.62% – 1.37%). For males, the proportion of visits were similar in 2018 and 2023, as the result of a decrease in 2021, followed by a subsequent increase in 2022. For race/Hispanic ethnicity, the proportion of substance use-related ED visits decreased for Non-Hispanic White and Asian patients from 2018 to 2023, and increased for Non-Hispanic Black, Hispanic, and Other race/ethnicity patients (Figure 2b).

**Figure 2.**
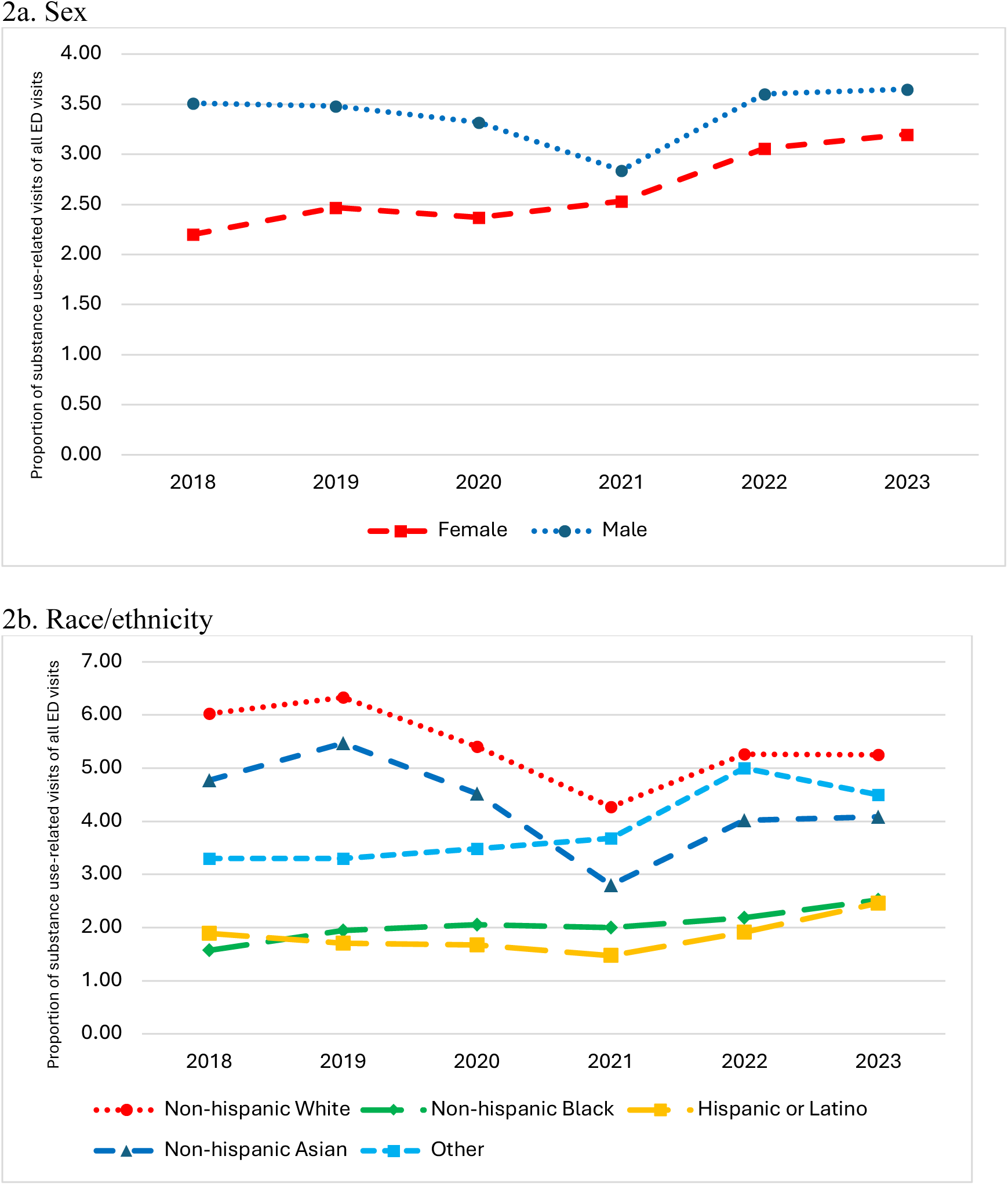
Substance use-related ED visits for youth by sex and race/ethnicity, 2018-2023.

### 3.4. Trends in substances involved in substance-use related visits

The three most common substances involved in substance use-related visits were alcohol (53.1%), cannabis (26.2%), and other psychoactive substances (17.5%) (Table 3). ED visits for alcohol decreased during the study period from 1.87% in 2018 to 1.55% in 2023 (Δ -0.33%, 95% CI -0.10% – -0.55%) (Figure 3a). Whereas, the proportion of ED visits for cannabis increased from 0.50% in 2018 to 1.20% in 2023 (Δ 0.70% (95% CI 0.55% – 0.86), with increases in ED visits across all age groups (p < 0.001) (Figures 3b-3d). Of substance use-related visits, the proportion of visits for cannabis increased from 17.9% of visits in 2018 to 35.3% in 2023 (Δ 17.4%, 95% CI 8.8% - 26.1%), with an increase in visits for cannabis for females (5.4% to 20.0% (Δ14.6%, 95% CI 5.1% - 24.0%)). Cannabis was the most common substance involved in substance-related visits for youth 12-14 years old, with cannabis-related ED visits increasing 0.85% (95% CI 0.58% - 1.12%) from 0.11% in 2018 to 0.95% in 2023. Visits for opioids, stimulants, and sedatives were less common, though there was an increase in visits for opioids during the study period (0.05% in 2018 to 0.12% in 2023 ((Δ 0.07%, 95% CI 0.02% - 0.12%)).

**Figure 3.**
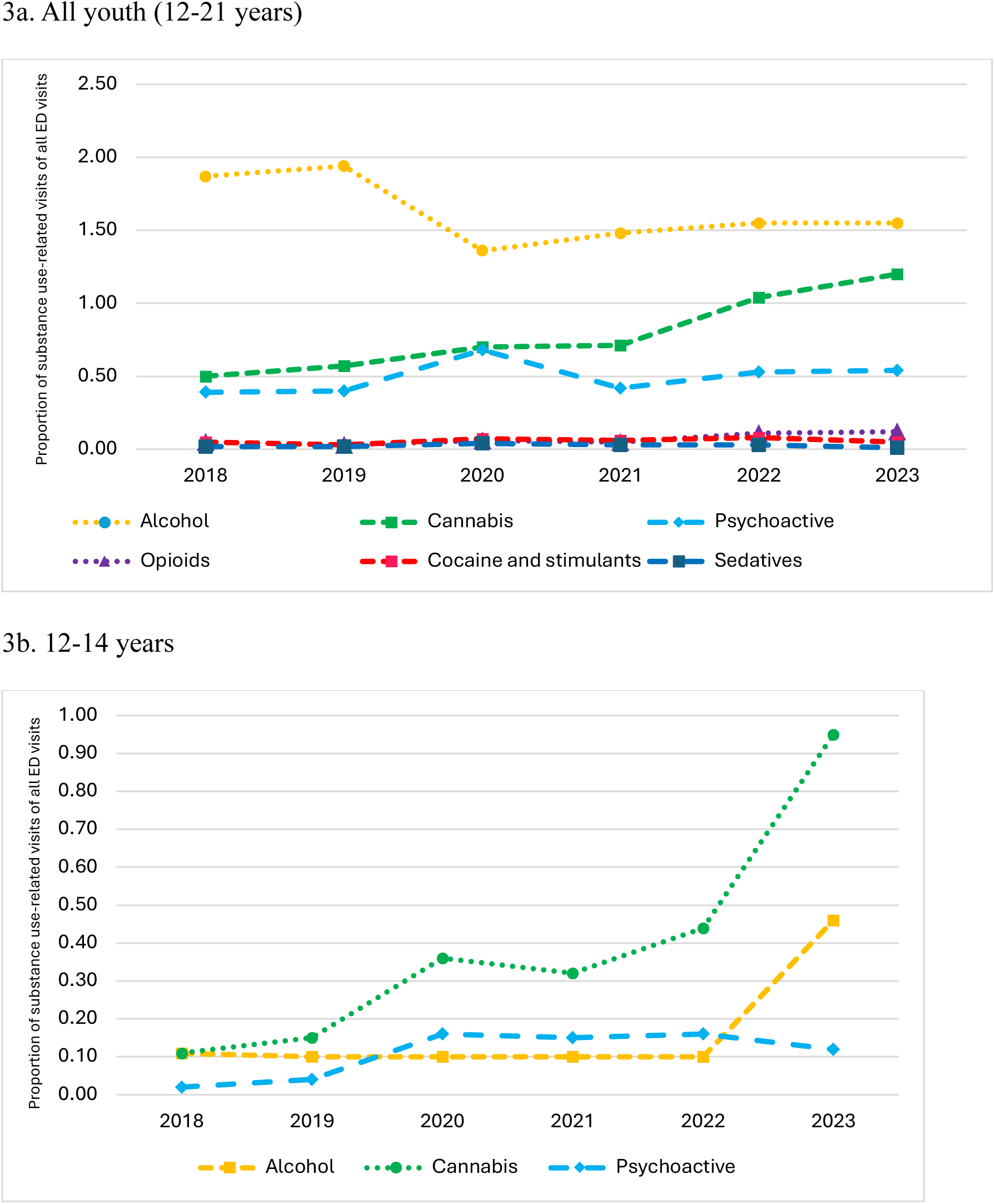
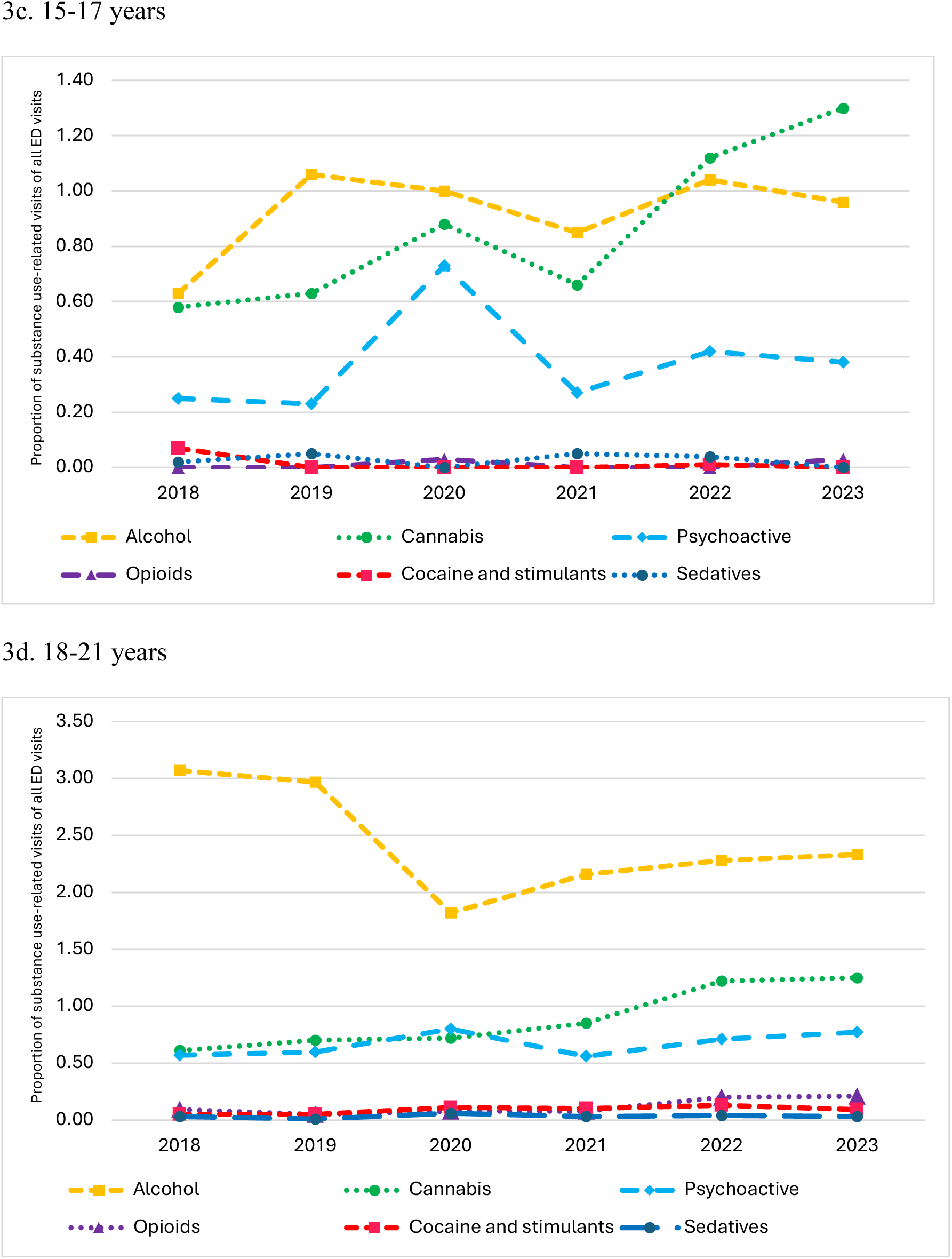
Substance use-related ED visits for youth by substance type and age, 2018-2023.

**Table 3.**
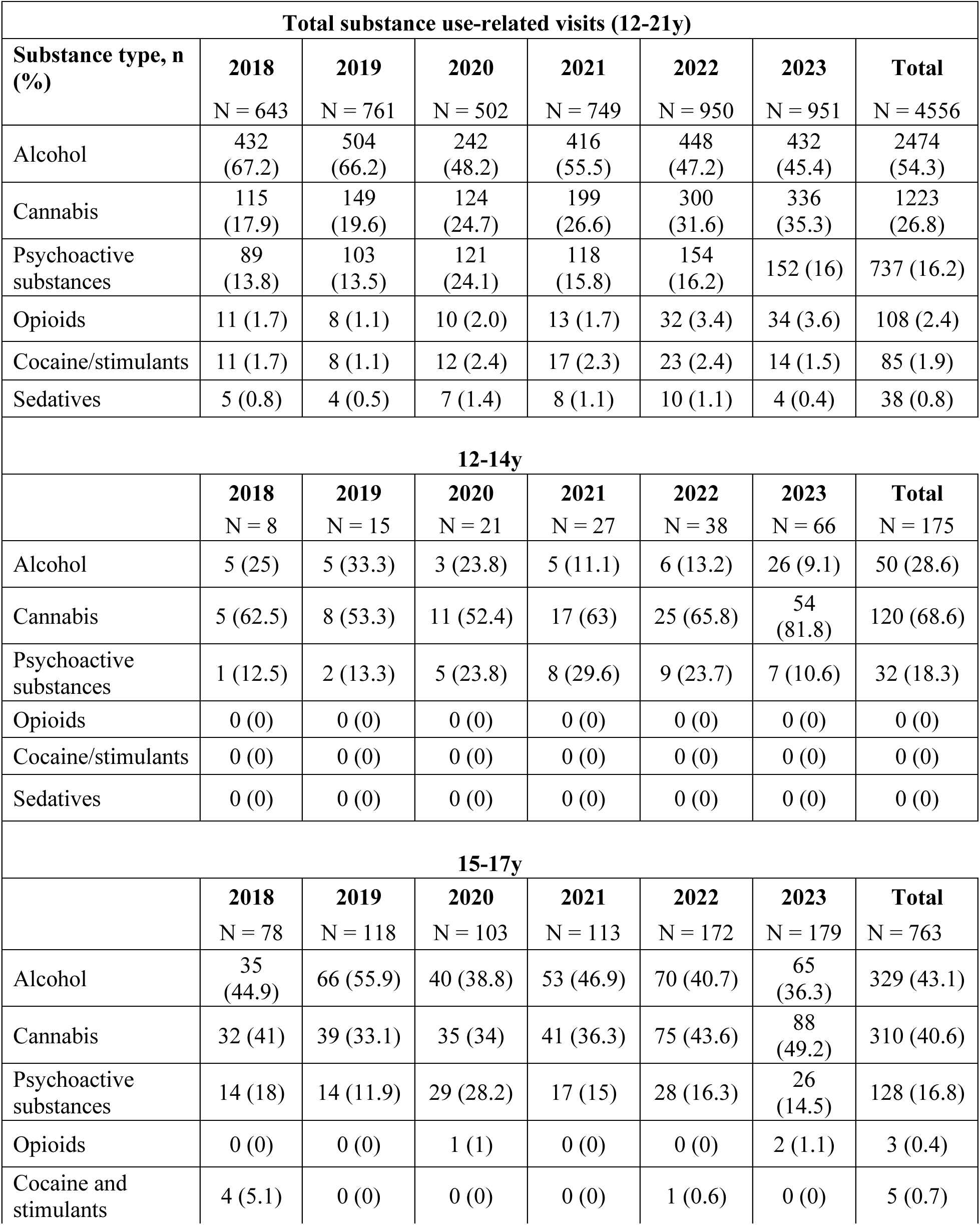
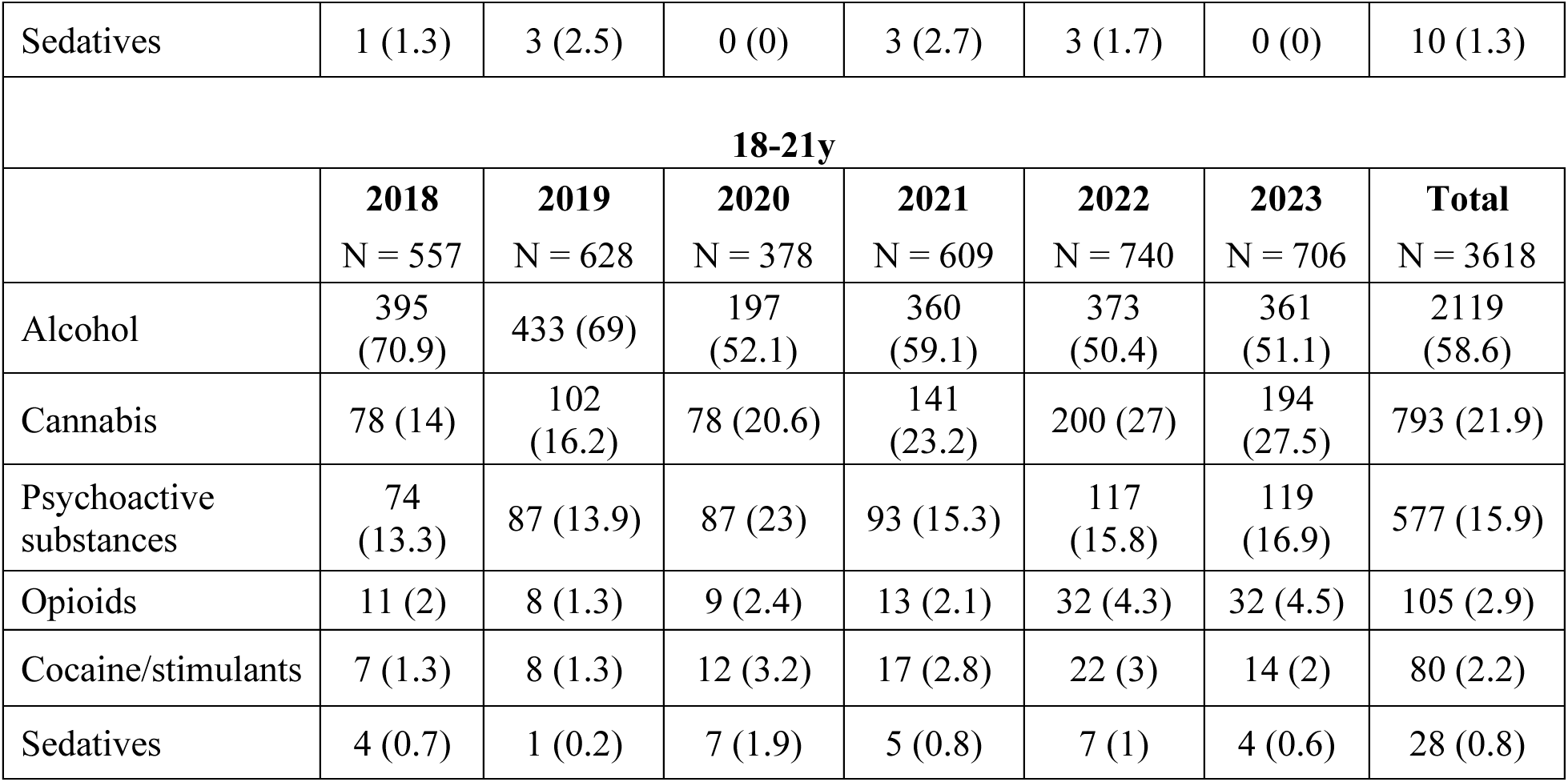
Substance use-related emergency department visits, by substance type and age group, 2018-2023.

### 3.5. ED visit disposition and revisits

The vast majority of ED visits for substance use had a disposition of discharge home (93.2%). 3.2% of visits resulted in an admission to the hospital, in 0.86% of visits the patient was transferred to psychiatry, and 1.9% of visits had a self-directed/premature discharge. There was an increase in admissions and self-directed discharges in 2020, but the dispositions were otherwise similar across the study period (Supplementary Material, Figure S1). Substance use-related visits for male patients had an increased odds of admission and compared to visits for alcohol, visits for all other substance types had an increased odds of admission (Supplementary Material, Table S2). In regard to ED revisits, 2.9% of visits involved patients that had an ED revisit for a substance use-related diagnosis within one week and 19.2% of visits involved patients had a revisit within one year. ED revisits within one week for substance use increased from 1.71 % in 2018 to 3.47% in 2023 (Supplementary Material, Figure S2). One-year revisits during the study period were similar over time. Males had increased odds of revisits at one week, revisits within one year were more likely for older youth, and compared to Hispanic patients, Non-Hispanic White, Asian, and Other race/ethnicity patients had a decreased odds of revisits at one week and one year (Supplementary Material, Table S3 and Table S4). Compared to alcohol, ED revisits within one week and one year were more likely for all other substance types, except for revisits within one week for sedatives.

## 4. Discussion

In this study of ED visits in an urban health care system, we found that substance use-related ED visits for youth increased from 2018 to 2023. Although males and older youth were more likely to have a substance use-related ED visit, the proportion of substance use-related ED visits increased most notably for younger adolescents and females, and for visits related to cannabis use. The majority of patients were discharged home from the ED, but ED revisits for substance use-related diagnoses were common.

The increase in substance use-related ED visits in our health system is substantial and most marked in recent years. A prior study found that substance use visits accounted for 2.1% of all ED visits in the US from 1997 to 2010,^7^ whereas in our health system, substance use-related ED visits were 3.4% of all ED visits for 12–21-year-olds by 2023. Our findings add to prior research that demonstrated an increase in ED visits for substance use in the first two years of the COVID-19 pandemic.^8^ The increase in ED visits is in contrast to national data reporting lower rates of drug use for adolescents in the US since the COVID-19 pandemic.^1,12^ These findings may be due to potential under-reporting in national self-reported surveys of substance use in youth, as well as higher risk substance use practices in youth leading to an increase in substance use-related problems, and/or the increased risk of the current illicit drug supply, which could lead to an increase in visits for emergent care.^13,14^

Of importance, ED visits increased markedly for younger youth (12-14 and 15-17 years old). In addition to the negative impact on neurocognitive development, the initiation of substance use in younger adolescents is associated with an increased risk of developing a substance use disorder (SUD).^3,4,15^ Substance use in adolescence is also associated with other high-risk behaviors.^16,17^ Given the rise in ED visits for younger adolescents, our findings highlight the need for ED interventions that target both older and younger youth. While there is considerable research on ED interventions for substance use in adults,^18-20^ there is limited research in youth. Most studies in youth have focused on brief, motivationally-based interventions for alcohol, with fewer studies on cannabis and opioids.^21-23^ Brief interventions in youth are feasible, acceptable, and may decrease substance use.^24-26^ Further research in adolescents, including younger adolescents, and across different substance types is warranted to determine the most effective and scalable ED interventions for youth. In addition, the vast majority of youth with SUDs do not receive treatment,^27^ and early intervention for SUDs may lead to more successful treatment;^28^ therefore, ED interventions should prioritize linkage to outpatient treatment.

Alcohol and cannabis were the most common substance types involved in substance-related ED visits in our healthcare system, which is consistent with national data on substances most commonly used by youth and prior literature on ED visits for substance use.^1,12^ ED visits for alcohol decreased during the study period, while visits for cannabis significantly increased across all age groups. The increase in ED visits for cannabis might be related to increased access to and availability of cannabis in the setting of changing cannabis legislation/legalization,^29,30^ as well as youth perception that using cannabis is not harmful.^1,31^ In addition to recreational cannabis use, youth may have unintentional exposures to cannabis due to copycat and lookalike edible cannabis products.^32^ For 12–14-year-olds in our study, cannabis was the most common substance involved in substance use-related ED visits, with a notable 764% relative increase in visits for cannabis during the study period. Adolescent cannabis use can lead to cannabis use disorder and may be associated with other morbidities, including psychosis, schizophrenia, and decreases in intelligent quotient as an adult,^4,33,34^ highlighting the need for targeted youth interventions for cannabis use.

Though opioid-related substance use visits were less common, during the study period opioid-related visits increased relatively by 140%. These visits were predominately for patients 18-21 years old. In light of the ongoing opioid epidemic and continued risks of the current illicit drug supply,^14^ these findings emphasize the importance of the ED visit as an opportunity for interventions including harm reduction, including naloxone distribution, and initiation of treatment with medications for opioid use disorder, as well as linkage to outpatient care.

During the study period, there were variations by sex and race/Hispanic ethnicity. Though males had a greater proportion of substance use-related ED visits throughout the study period, visits for females increased significantly. This is similar to prior studies and may be reflective of worsening mental health in female youth in recent years.^8,35^ The proportion of substance related ED visits for Non-Hispanic Black, Hispanic, and Other race/ethnicity patients increased from 2018 to 2023. Disproportionate increases in substance use visits for Non-Hispanic Black and Hispanic youth has been demonstrated in prior studies.^8,36^ Future research should investigate the underlying cause for these increasing trends and ways to ensure equitable care for youth with substance use.

The majority of patients were discharged home and revisits to the ED for substance use were common, with one in five visits involving patients that returned to the ED for a substance use-related diagnosis within one year and a smaller proportion within one week. Our findings on ED revisits are similar to a study in Ontario, Canada, which found that the proportion of repeated ED visits (within one year) for substance use was 20.2% in 2018 and males were more likely to have revisits to the ED.^37^ These findings highlight the importance of brief interventions in the ED, as this is a critical health care access point for these youth (that per our findings are rarely admitted to the hospital), and the development of ED interventions that target linkage to outpatient care. Though there is limited research on linkage to care for adolescents with substance use, research on adults strongly supports ED linkage to care.^38-40^ Future studies may aid in guiding best practices for youth in the ED and determine if follow up care after an ED visit prevents morbidity and mortality from substance use, and in turn, may prevent patients from returning to the ED for similar substance use-related harms, illnesses, and/or injuries.

There are several limitations to our study. First, the study was contingent on ED visits for substance use identified using diagnostic ICD-10-CM coding, which could be subject to inaccuracies in classification. Our findings could underestimate substance use-related ED visits, because not all visits for substance use may be coded as such. Second, the data from this study comes from an urban health system, and while it is a multicenter health system, our findings may not be externally valid to all ED settings and locations. Third, our small sample size for ED visits for substance use for certain age groups and race/Hispanic ethnicity categories within our population may limit our ability to make definitive conclusions based on our analysis. Future larger studies are needed to further examine trends in these subsets of the population. Finally, due to very few nicotine-related ED visits during the study period (n = 12), we did not include nicotine as a substance type for substance related-ED visits. While other studies have identified nicotine as a common substance involved in ED visits, this was not the case for our health system.

## 5. Conclusion

Substance use-related ED visits for youth increased in this urban health care system from 2018 to 2023, with notable increases in visits for younger age groups (12-14 years and 15-17 years). Alcohol and cannabis were the most common substances involved, with an increase in visits for cannabis over time across all age groups. Revisits for substance use were common. These findings can inform targeted ED-based interventions for substance use in youth.

## Conflicts of Interest

All authors declare that they have no conflict of interest.

## Prior presentation of data

Preliminary data from this manuscript was previously presented at the Pediatric Academic Society Meeting (Washington, D.C., 2023).

## Sources of funding

Roland C. Merchant was supported by a National Institute on Drug Abuse career mentoring award (K24DA044858).

## Data Availability

Research data includes sensitive or confidential information such as patient data so is not available.

## Notes

### Competing Interest Statement

The authors have declared no competing interest.

### Author Declarations

The IRB of the Icahn School of Medicine at Mount Sinai gave ethical approval for this work.

## References

1. Miech RA, Johnston LD, Patrick ME, O’Malley PM, Bachman JG. Monitoring the Future National Survey Results on Drug Use, 1975–2023: Secondary school students. 2023. https://monitoringthefuture.org/results/annual-reports/

2. Friedman J, Godvin M, Shover CL, Gone JP, Hansen H, Schriger DL. Trends in Drug Overdose Deaths Among US Adolescents, January 2010 to June 2021. JAMA. 2022;327(14):1398-1400. doi:10.1001/jama.2022.2847

3. Volkow ND, Han B, Einstein EB, Compton WM. Prevalence of Substance Use Disorders by Time Since First Substance Use Among Young People in the US. JAMA Pediatrics. 2021;175(6):640–643. doi:10.1001/jamapediatrics.2020.6981

4. Han B, Compton WM, Blanco C, Jones CM. Time since first cannabis use and 12-month prevalence of cannabis use disorder among youth and emerging adults in the United States. Addiction. 2019;114(4):698–707. 10.1111/add.14511

5. Centers for Disease Control and Prevention. National Centers for Injury Prevention and Control. Web-based Injury Statistics Query and Reporting System (WISQARS) [online]. 2020. Accessed March 10 2023.

6. Ziv A, Boulet JR, Slap GB. Emergency department utilization by adolescents in the United States. Pediatrics. Jun 1998;101(6):987–94. doi:10.1542/peds.101.6.987

7. Fahimi J, Aurrecoechea A, Anderson E, Herring A, Alter H. Substance abuse and mental health visits among adolescents presenting to US emergency departments. Pediatr Emerg Care. May 2015;31(5):331–8. doi:10.1097/pec.0000000000000421

8. Ball A, Hadland S, Rodean J, Hall M, Mendoza J, Ahrens K. Trends in Substance-Related Visits Among Youth to US Children’s Hospitals, 2016-2021: An Analysis of the Pediatric Health Information System Database. J Adolesc Health. Mar 25 2024;doi:10.1016/j.jadohealth.2024.02.016

9. Bommersbach TJ, McKean AJ, Olfson M, Rhee TG. National Trends in Mental Health– Related Emergency Department Visits Among Youth, 2011-2020. JAMA. 2023;329(17):1469-1477. doi:10.1001/jama.2023.4809

10. Lo CB, Bridge JA, Shi J, Ludwig L, Stanley RM. Children’s Mental Health Emergency Department Visits: 2007–2016. Pediatrics. 2020;145(6) doi:10.1542/peds.2019-1536

11. Centers for Disease Control and Prevention. National Center for Health Statistics - ICD- 10-CM. Accessed July 10, 2022, https://www.cdc.gov/nchs/icd/icd-10-cm/?CDC_AAref_Val=https://www.cdc.gov/nchs/icd/icd-10-cm.htm

12. Substance Abuse and Mental Health Services Administration. Key substance use and mental health indicators in the United States: Results from the 2022 National Survey on Drug Use and Health (HHS Publication No. PEP23-07-01-006, NSDUH Series H-58). 2023.

13. Roehler DR, Olsen EO, Mustaquim D, Vivolo-Kantor AM. Suspected nonfatal drug-related overdoses among youth in the US: 2016-2019. Article. Pediatrics. 2021;147(1)e2020003491. doi:10.1542/PEDS.2020-003491

14. Tanz LJ, Dinwiddie AT, Mattson CL, O’Donnell J, Davis NL. Drug Overdose Deaths Among Persons Aged 10–19 Years — United States, July 2019–December 2021. Journal Article. MMWR Morb Mortal Wkly Rep 71(50):1576-1582. 2022;71(50)

15. Hanson KL, Medina KL, Padula CB, Tapert SF, Brown SA. Impact of Adolescent Alcohol and Drug Use on Neuropsychological Functioning in Young Adulthood: 10-Year Outcomes. J Child Adolesc Subst Abuse. Jan 1 2011;20(2):135–154. doi:10.1080/1067828x.2011.555272

16. Clayton HB, Bohm MK, Lowry R, Ashley C, Ethier KA. Prescription Opioid Misuse Associated With Risk Behaviors Among Adolescents. Am J Prev Med. Oct 2019;57(4):533–539. doi:10.1016/j.amepre.2019.05.017

17. Centers for Disease Control and Prevention. Youth Risk Behavior Survey Data Summary & Trends Report, 2009–2019. Atlanta, GA: U.S. Department of Health and Human Services, Centers for Disease Control and Prevention, Office of Infectious Diseases, NCHHSTP; 2020. 2020.

18. Hawk K, D’Onofrio G. Emergency department screening and interventions for substance use disorders. Addict Sci Clin Pract. Aug 6 2018;13(1):18. doi:10.1186/s13722-018-0117-1

19. Blow F, Walton M, Bohnert A, et al. A Randomized Controlled Trial of Brief Interventions to Reduce Drug Use Among Adults in a Low Income Urban Emergency Department: The Healthi ER You Study: Drug BIs Among Adults in an ED. Addiction. 01/26 2017;112doi:10.1111/add.13773

20. Welch AE, Jeffers A, Allen B, Paone D, Kunins HV. Relay: A Peer-Delivered Emergency Department–Based Response to Nonfatal Opioid Overdose. American Journal of Public Health. 2019;109(10):1392–1395. doi:10.2105/ajph.2019.305202

21. Bernstein J, Heeren T, Edward E, et al. A brief motivational interview in a pediatric emergency department, plus 10-day telephone follow-up, increases attempts to quit drinking among youth and young adults who screen positive for problematic drinking. Acad Emerg Med. Aug 2010;17(8):890–902. doi:10.1111/j.1553-2712.2010.00818.x

22. Bernstein E, Edwards E, Dorfman D, Heeren T, Bliss C, Bernstein J. Screening and brief intervention to reduce marijuana use among youth and young adults in a pediatric emergency department. Acad Emerg Med. Nov 2009;16(11):1174–85. doi:10.1111/j.1553-2712.2009.00490.x

23. Walton MA, Chermack ST, Blow FC, et al. Components of Brief Alcohol Interventions for Youth in the Emergency Department. Substance abuse. 2015;36(3):339–349. doi:10.1080/08897077.2014.958607

24. Spirito A, Monti PM, Barnett NP, et al. A randomized clinical trial of a brief motivational intervention for alcohol-positive adolescents treated in an emergency department. J Pediatr. Sep 2004;145(3):396–402. doi:10.1016/j.jpeds.2004.04.057

25. Arnaud N, Diestelkamp S, Wartberg L, Sack PM, Daubmann A, Thomasius R. Short- to Midterm Effectiveness of a Brief Motivational Intervention to Reduce Alcohol Use and Related Problems for Alcohol Intoxicated Children and Adolescents in Pediatric Emergency Departments: A Randomized Controlled Trial. Acad Emerg Med. Feb 2017;24(2):186–200. doi:10.1111/acem.13126

26. Cunningham RM, Chermack ST, Ehrlich PF, et al. Alcohol Interventions Among Underage Drinkers in the ED: A Randomized Controlled Trial. Pediatrics. 2015;136(4):e783–e793. doi:10.1542/peds.2015-1260

27. Substance Abuse and Mental Health Services Administration. Key substance use and mental health indicators in the United States: Results from the 2021 National Survey on Drug Use and Health (HHS Publication No. PEP22-07-01-005, NSDUH Series H-57). 2022.

28. Dennis ML, Scott CK, Funk R, Foss MA. The duration and correlates of addiction and treatment careers. J Subst Abuse Treat. 2005;28 Suppl 1:S51–62. doi:10.1016/j.jsat.2004.10.013

29. Wang GS, Davies SD, Halmo LS, Sass A, Mistry RD. Impact of Marijuana Legalization in Colorado on Adolescent Emergency and Urgent Care Visits. J Adolesc Health. Aug 2018;63(2):239–241. doi:10.1016/j.jadohealth.2017.12.010

30. Yeung MEM, Weaver CG, Hartmann R, Haines-Saah R, Lang E. Emergency Department Pediatric Visits in Alberta for Cannabis After Legalization. Pediatrics. Oct 2021;148(4)doi:10.1542/peds.2020-045922

31. Harrison ME, Kanbur N, Canton K, et al. Adolescents’ Cannabis Knowledge and Risk Perception: A Systematic Review. Journal of Adolescent Health. 2024/03/01/ 2024;74(3):402–440. 10.1016/j.jadohealth.2023.09.014

32. Ompad DC, Snyder KM, Sandh S, et al. Copycat and lookalike edible cannabis product packaging in the United States. Drug Alcohol Depend. Jun 1 2022;235:109409. doi:10.1016/j.drugalcdep.2022.109409

33. Levine A, Clemenza K, Rynn M, Lieberman J. Evidence for the Risks and Consequences of Adolescent Cannabis Exposure. Journal of the American Academy of Child & Adolescent Psychiatry. 2017/03/01/ 2017;56(3):214-225. 10.1016/j.jaac.2016.12.014

34. Meier MH, Caspi A, Ambler A, et al. Persistent cannabis users show neuropsychological decline from childhood to midlife. Proc Natl Acad Sci U S A. Oct 2 2012;109(40):E2657–64. doi:10.1073/pnas.1206820109

35. Krass P, Dalton E, Doupnik SK, Esposito J. US Pediatric Emergency Department Visits for Mental Health Conditions During the COVID-19 Pandemic. JAMA Network Open. 2021;4(4):e218533–e218533. doi:10.1001/jamanetworkopen.2021.8533

36. Masonbrink AR, Middlebrooks L, Gooding HC, et al. Substance Use Disorder Visits Among Adolescents at Children’s Hospitals During COVID-19. Journal of Adolescent Health. 2022/04/01/ 2022;70(4):673-676. 10.1016/j.jadohealth.2021.12.024

37. Kim S, Weekes J, Young MM, Adams N, Kolla NJ. Trends of repeated emergency department visits among adolescents and young adults for substance use: A repeated cross-sectional study. PLOS ONE. 2023;18(2):e0282056. doi:10.1371/journal.pone.0282056

38. Kmiec J, Suffoletto B. Implementations of a text-message intervention to increase linkage from the emergency department to outpatient treatment for substance use disorders. Journal of Substance Abuse Treatment. 2019/05/01/ 2019;100:39-44. 10.1016/j.jsat.2019.02.005

39. D’Onofrio G, Degutis LC. Integrating Project ASSERT: a screening, intervention, and referral to treatment program for unhealthy alcohol and drug use into an urban emergency department. Acad Emerg Med. Aug 2010;17(8):903–11. doi:10.1111/j.1553-2712.2010.00824.x

40. D’Onofrio G, O’Connor PG, Pantalon MV, et al. Emergency Department–Initiated Buprenorphine/Naloxone Treatment for Opioid Dependence: A Randomized Clinical Trial. JAMA. 2015;313(16):1636–1644. doi:10.1001/jama.2015.3474

