## Supplementary Material for "Trends in Substance Use-related Emergency Department Visits by Youth, 2018-2023"

**Table S1. ICD-10-CM codes for substance use-related emergency department visits**

| **Substance** | **ICD-10-CM Codes** |
| --- | --- |
| Alcohol | F10.1X, F10.2X, F10.9X |
| Cannabis | F12.1X, F12.2X, F12.9X |
| Opioids | F11.1X, F11.2X, F11.9X |
| Cocaine | F14.1X, F14.2X, F14.9X |
| Stimulants | F15.1X, F15.2X, F15.9X |
| Hallucinogens | F16.1X, F16.2X, F16.9X |
| Sedatives | F13.1X. F13.2X, F13.9X |
| Inhalants | F18.1X, F18.2X, F18.9X |
| Other Psychoactives | F19.1X, F19.2X, F19.9X |

**Table S2. Factors associated with a substance use-related ED visit disposition of admission to the hospital**

|  | **Adjusted Odds Ratio (95% CI)** |
| --- | --- |
| **Age** | |
| 12-14 years  15-17 years  18-21 years | Reference  3.13 (0.39 – 24.74)  6.32 (0.84 – 47.80) |
| **Sex** | |
| Female  Male | Reference  1.87 (1.26 – 2.78) |
| **Race/Ethnicity** | |
| Hispanic  Non-Hispanic White  Non-Hispanic Black  Asian  Other | Reference  0.87 (0.49 – 1.56)  0.97 (0.54 –1.73)  1.81 (0.83 – 3.92)  1.34 (0.81 – 2.22) |
| **Substance Use Type** | |
| Alcohol  Cannabis  Psychoactive substances  Opioids  Cocaine/stimulants  Sedatives | Reference  3.21 (1.94 – 5.31)  3.01 (1.77 – 5.10)  6.26 (2.85 – 13.75)  9.42 (4.38 – 20.25)  32.38 (14.63 – 71.68) |

**Table S3. Factors associated with substance use-related ED revisits visits within one week**

|  | **Adjusted Odds Ratio (95% CI)** |
| --- | --- |
| **Age** | |
| 12-14 years  15-17 years  18-21 years | Reference  1.05 (0.34 – 3.24)  2.06 (0.74 – 5.71) |
| **Sex** | |
| Female  Male | Reference  1.47 (1.02 – 2.12) |
| **Race/Ethnicity** | |
| Hispanic  Non-Hispanic White  Non-Hispanic Black  Asian  Other | Reference  0.38 (0.22 – 0.66)  1.14 (0.74 –1.76)  0.29 (0.09 – 0.94)  0.35 (0.20 –0.59) |
| **Substance Use Type** | |
| Alcohol  Cannabis  Psychoactive substances  Opioids  Cocaine/stimulants  Sedatives | Reference  2.83 (1.69 – 4.73)  3.49 (2.07 – 5.88)  7.37 (3.53 – 15.38)  6.12 (2.57 –14.56)  2.21 (0.29 – 17.02) |

**Table S4. Factors associated with substance use-related ED revisits within one year**

|  | **Adjusted Odds Ratio (95% CI)** |
| --- | --- |
| **Age** | |
| 12-14 years  15-17 years  18-21 years | Reference  1.18 (0.77 – 1.80)  1.55 (1.05 – 2.29) |
| **Sex** | |
| Female  Male | Reference  1.10 (0.94 – 1.29) |
| **Race/Ethnicity** | |
| Hispanic  Non-Hispanic White  Non-Hispanic Black  Asian  Other | Reference  0.45 (0.36 – 0.57)  1.00 (0.81 –1.24)  0.45 (0.31 – 0.65)  0.28 (0.22 – 0.35) |
| **Substance Use Type** | |
| Alcohol  Cannabis  Psychoactive substances  Opioids  Cocaine/stimulants  Sedatives | Reference  2.65 (2.18 – 3.23)  2.70 (2.17 – 3.37)  4.49 (2.88 – 6.99)  4.30 (2.64 – 7.00)  4.31 (2.19 – 8.50) |

**Figure S1. ED disposition for substance use-related visits, 2018-2023**

Proportion of substance use-related ED visits

|  | Discharge | Admission | Premature discharge | Transfer to psych |
| --- | --- | --- | --- | --- |
| 2018 | 94.4% | 4.16% | 1.12% | 0.32% |
| 2019 | 95.11% | 2.25% | 1.59% | 1.06% |
| 2020 | 89.52% | 5.24% | 3.43% | 1.81% |
| 2021 | 93.96% | 3.36% | 2.15% | 0.54% |
| 2022 | 95.11% | 2.34% | 1.81% | 0.74% |
| 2023 | 94.18% | 2.96% | 1.9% | 0.95% |

**Figure S2. ED revisits for substance use-related diagnoses, 2018-2023**

Proportion of substance use-related ED visits

|  | Revisits within a week | Revisits within one year |
| --- | --- | --- |
| 2018 | 1.71 | 18.66 |
| 2019 | 1.97 | 16.16 |
| 2020 | 3.78 | 25.1 |
| 2021 | 3.34 | 19.09 |
| 2022 | 3.37 | 21.89 |
| 2023 | 3.47 | 16.4 |
